# Gyroscope Vector Magnitude: A proposed measure for accurately measuring angular velocities

**DOI:** 10.1101/2022.10.05.22280752

**Authors:** Howard Chen, Mark Schall, Nathan Fethke

## Abstract

High movement velocities are among the primary risk factors for work-related musculoskeletal disorders (MSDs). Ergonomists have commonly used two methods to calculate angular movement velocities of the upper arms using inertial measurement units (accelerometers and gyroscopes). Generalized velocity is the speed of movement traveled on the unit sphere per unit time. Inclination velocity is the derivative of the postural inclination angle relative to gravity with respect to time. Neither method captures the full extent of upper arm angular velocity. We propose a new method, the gyroscope vector magnitude (GVM), and demonstrate how GVM captures angular velocities around all motion axes and more accurately represents the true angular velocities of the upper arm. We use optical motion capture data to demonstrate that the previous methods for calculating angular velocities capture 89% and 77% relative to our proposed method. We propose GVM as the standard metric for reporting angular arm velocities in future research.

## Introduction

Inertial measurement units (IMUs) can quantify occupational risk factors for musculoskeletal disorders (MSDs), such as high movement velocities (Arvidsson et al., 2021; Kersten and Fethke, 2019; Schall et al., 2021). An IMU contains triaxial gyroscopes, accelerometers, and magnetometers, which allows a variety of approaches to kinematic measurements such as full-body inertial motion capture (Robert-Lachaine et al., 2019), inclinometry using gyroscope and accelerometer measurements (IMU-based inclinometer) (Chen et al., 2020; Fan et al., 2021; Schall et al., 2021), and inclinometry using accelerometer measurements (accelerometer-based inclinometer) (Chen et al., 2018). In contrast to full-body inertial motion capture, IMU-based inclinometers are not susceptible to errors from gyroscopic drift or ambient magnetic field disturbances (Chen et al., 2020; Schall Jr. et al., 2016b). However, the primary limitation of IMU-based inclinometers is an inability to measure movement around the gravity vector.

The angular velocities of body segments, particularly the upper arm, have been frequently quantified with inclinometers (Douphrate et al., 2012; Granzow et al., 2018; Schall Jr. et al., 2016a; Veiersted et al., 2008) using one of two computational methods: inclination velocity (Douphrate et al., 2012; Granzow et al., 2018; Schall et al., 2021) or generalized velocity (Arvidsson et al., 2012; Veiersted et al., 2008). Inclination velocity is obtained by derivation of upper arm inclination angle (relative to gravity) with respect to time. Generalized velocity is defined as the distance traveled on the unit sphere per time unit (Hansson et al., 2001) and, in general, produces higher velocity magnitudes than inclination velocity because rotations in multiple directions are factored in the calculation (Fan et al., 2021). Differences in measurements between the inclination and generalized velocity approaches have been highlighted (Fan et al., 2021; Forsman et al., 2022a) and are important given recently proposed occupational exposure thresholds for movement velocities (Arvidsson et al., 2021). Inclination and generalized velocity can be calculated using accelerometer data alone, for which motion introduces measurement error (Chen et al., 2018), or in combination with gyroscope data and a sensor fusion algorithm (Fan et al., 2021), which reduces measurement error but increases computational overhead. Forsman et al. (2020b) proposed conversion models to resolve differences between accelerometer and IMU-derived upper arm inclination velocities and generalized velocities. They indicated that more studies are needed to determine one common standard metric for reporting upper arm angular velocities. One motivating factor is that neither inclination velocity nor generalized velocity capture angular velocity in all movement directions.

Few studies have evaluated the accuracy of accelerometer and IMU-derived angular velocity measurements or their associated movement summary measures used for health-based decision-making in the context of occupational ergonomics (Chen et al., 2020, 2018; Schall Jr. et al., 2016b, 2015). We reported a root-mean-square (RMS) error of 79°/s for accelerometer-derived upper arm inclination velocity for ‘fast’ motion during a cyclic material handling task (45 cycles/min), using optical motion capture (OMC) as a reference (Chen et al., 2018). An important observation from that study was the underestimation of the extreme upper arm postures and velocities (i.e., 90th percentiles) at increased motion speeds. Such underestimation may impact observed associations between summary measures of exposure and musculoskeletal health outcomes in epidemiologic studies. Fan et al. (2021) observed an average difference of 50°/s between accelerometer- and IMU-derived generalized velocities, despite mean differences <2° in inclination angles. Notably, they observed that accelerometer-derived upper arm generalized velocities were 100°/s greater than the IMU-derived upper arm generalized velocities for the 90^th^ and 99^th^ percentile summary measures. To our knowledge, however, the accuracy of neither accelerometer-nor IMU-derived generalized velocities has been validated against OMC.

Because of the availability of gyroscope data from an IMU, it seems counterintuitive to rely on accelerometer data in addition to gyroscope data when calculating upper arm angular velocity. The gyroscope vector magnitude incorporates rotation in all movement directions, in contrast to inclination and generalized velocity, and does not impose a computational penalty of using a sensor fusion algorithm. The gyroscope vector magnitudes from wrist- and hip-worn IMUs have been used to estimate energy expenditure during activities of daily living (Hibbing et al., 2018; Marcotte et al., 2018). In an ergonomics context, Manivasagam and Yang (2022) reported the difference in gyroscope vector magnitudes from IMUs positioned on the hand and forearm as a measure of “total” wrist angular velocity (vs. velocity obtained through differentiation of data from a standard biaxial electrogoniometer). However, Manivasagam and Yang (2022) did not provide a biomechanical definition for this quantity, which is necessary for researchers and practitioners to interpret its meaning.

In this paper, we (i) expand the work of Fan et al. (2021) by evaluating the accuracy of accelerometer and IMU-derived generalized velocity measurements for the upper arm and their associated summary metrics, and (ii) expand on the work of Manivasgam and Yang (2022)by providing a biomechanical definition of gyroscope vector magnitude necessary for its use as an exposure measure in occupational ergonomics applications. We also use OMC measurements to demonstrate how gyroscope vector magnitude captures angular velocity to a fuller extent than both inclination and generalized velocity.

## Methods

### Angular Velocity Calculation Methods

#### Inclination Velocity

We define inclination velocity as the absolute value of the rate of change of upper arm elevation angle (relative to gravity) with respect to time. Inclination velocity (incVel) can be calculated as follows:

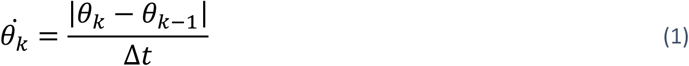

where θ_*k*_ and θ_*k-1*_ are the upper arm elevation angles relative to gravity at instance *k* and *k* − 1, respectively, and Δ*t* is the sampling period.

Upper arm elevation is calculated as follows:

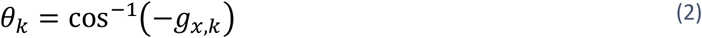

where *g*_*x,k*_ is the x-component of the normalized gravitational vector in the sensor frame. Note that 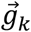 is the sensor frame. Therefore, this value will change with sensor inclination. 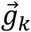 can therefore be thought of as the normalized accelerometer measurements in the absence of non-gravitational acceleration. Since 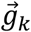 can be decomposed into ‘pitch’ and roll’ angles (Pedley, 2013), 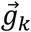 canalso be considered as the combination of ‘pitch’ and ‘roll’ angles. Equation (2) assumes that the x-axis of the IMU is aligned with the upper arm, with positive x oriented distally. While this method is easily interpretable, incVel does not change when the upper arm is moved purely around gravity, i.e., upper arm movements with no change in elevation angle.

#### Generalized Velocity

Generalized velocity has been defined as the angle traveled on the unit sphere per time unit (Fan et al., 2021). Detailed explanations of generalized velocities can be found elsewhere (Fan et al., 2021; Hansson et al., 2001, 2006). In contrast to incVel, this approach considers two movement directions, capturing angular velocities to a fuller extent than incVel.

Generalized velocity can be calculated as follows (Fan et al., 2021):

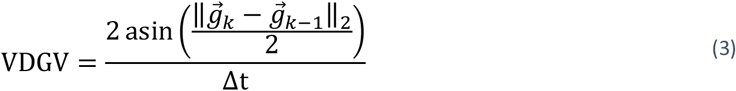

where 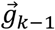 and 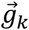 are the normalized gravitational vectors in the sensor frame at instances k-1 and k, respectively. Since this method requires differencing the gravitational vectors, we will refer to this calculation method as vector-differenced generalized velocity (VDGV).

The primary difference between incVel and VDGV can be illustrated as follows: if we assume that the IMU is attached to the upper arm such that the sensor z-axis represents vertical (i.e., the plane of elevation), the y-axis represents flexion/extension, and the x-axis represents abduction/adduction, velocity due to pure flexion/extension will register the same value for both incVel and VDGV. Pure abduction/adduction motion will register on VDGV but not incVel. Pure plane of elevation motion will register on neither.

The angle (*β*) between two unit vectors can be calculated by taking the arc-cosine of the dot product. Given 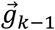 and 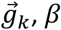 *β* can also be conceptualized as the net angular change between successive unit vectors (*4*). Therefore, dividing *β* by the sample period yields the rate of angular change between successive directional unit vectors, or the angular velocity 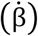 (*5*). This provides the same quantity as (*3*).

VDGV can, therefore, be alternatively calculated as follows:

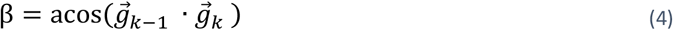

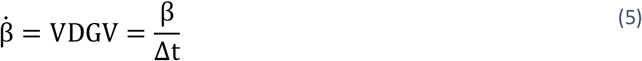

where · is the vector dot product.

There are two fundamental limitations to using incVel or VDGV for quantifying angular velocities. First, when 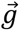 is derived from accelerometer data alone, the accuracy of incVel is adversely affected by non-gravitational acceleration, particularly during high-speed movements (Chen et al., 2020). The measurement accuracy of incVel can be improved, however, by combining accelerometer and gyroscope measurements with a sensor fusion algorithm (e.g., Kalman filter or complementary filter) (Chen et al., 2020, 2018; Johnson et al., 2022; Ligorio and Sabatini, 2015). Second, neither incVel nor VDGV incorporates rotations around gravity; therefore, neither method captures the full extent of angular velocity.

#### Gyroscope Vector Magnitude

We propose using the gyroscope vector magnitude (GVM) to quantify movement velocities rather than incVel or VDGV. GVM can capture angular velocity around all movement directions since obtaining 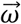 by differencing orientation measurements will present no further loss of information beyond its initial orientation. This can be verified by calculating 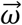 by differencing orientation measurements and subsequently re-calculating orientation using 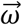, Δ*t*, and an initial orientation (Appendix B). The result will match the original set of orientation measurements.

GVM is calculated as follows:

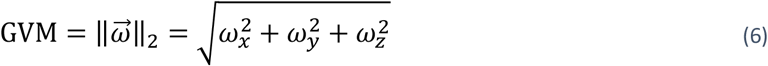

where *ω*_*x*_, *ω*_*y*_, and *ω*_*z*_ are the angular velocity measurements around the sensor x, y, and z-axes. The vector magnitude of 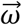 is defined as the rate of directional change between successive coordinate frames, and a mathematical derivation is shown in Appendix B. While 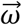 is most easily obtained directly from gyroscope measurements, it can also be obtained by differencing orientation measurements (See Appendix C).

### Experimental Method

#### Participants and Experimental Task

This study used data collected for a laboratory evaluation of IMUs among 13 right-handed participants (11 male; mean age= 27.2 ± 6.6 years). We refer to previous studies for detailed protocols and instrumentation (Chen et al., 2020, 2018, 2017). Briefly, exclusion criteria included any self-reported cases of (i) physician-diagnosed MSD in the previous six months, (ii) pain during the two weeks prior to study enrollment, and (iii) history of orthopedic surgery in the upper extremity (shoulder, elbow, wrist, hand). Written informed consent was obtained prior to data collection. The study protocol was approved by the University of Iowa Institutional Review Board.

Participants completed six one-minute trials of a repetitive reaching task that required transferring wooden dowels (2 cm diameter x 8 cm length) from a container placed directly in front of each participant’s waist to a container placed approximately 45° offset diagonally from the participant at shoulder level. Participants completed two trials at each of three transfer rates dictated by a metronome: slow (15 cycles/min), medium (30 cycles/min), and fast (45 cycles/min). Experimental conditions were randomized to control for potential order effects. Participants were acclimated to the assigned transfer rate before recording OMC measurements for each trial, followed by a five-minute rest period. This experiment used the material transfer rate as a proxy for movement speed.

#### Instrumentation

Participants were fitted with an IMU on the lateral aspect of the dominant upper arm midway between the acromion and the lateral epicondyle (Opal, APDM, Inc. Portland, OR; also sold as series SXT, Nexgen Ergonomics, Inc., Pointe Claire, Quebec, CA). The IMU was secured using Velcro® straps. Raw accelerometer and gyroscope measurements were sampled from the IMUs at 128 Hz. A six-camera OMC system (Optitrack Flex 13, NaturalPoint, Inc., Corvallis, OR, USA) sampling at 120 Hz tracked the position of four reflective markers attached rigidly to the surface of the IMU. Both the IMU and OMC systems were calibrated using manufacturer-specified procedures, including removing initial gyroscope bias using the manufacturer’s software. The OMC measurements were recorded for the duration of each trial (one minute). The IMU measurements were recorded for the entirety of each testing session (>30 minutes).

##### Optical Motion Capture

The OMC provides a quaternion 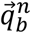 describing the spatial orientation of a given rigid body *b* relative to frame *n*. 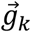 is obtained from OMC using (*19*) in Appendix C. Note that 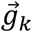 corresponds to the direction of gravity in the body frame (i.e., perfect accelerometer measurement), assuming that frame *n* is aligned with the gravity vector. Once 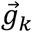 was obtained, incVel was calculated using (*1*) and (*2*), and VDGV was calculated using (*3*). These measurements will be referred to as omc-incVel and omc-VDGV. Further, 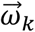 was calculated from OMC using (*20*) and will be referred to as omc-GVM. The three OMC-based measurements were used as reference signals with which to compare the sensor-based measurements.

##### Inertial Measurement Unit

The raw accelerometer data were first low-pass filtered (2nd order Butterworth, 3 Hz corner frequency). Then, 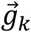 was calculated by normalizing the filtered accelerometer measurements before using (*1*) and (*2*) to calculate incVel (acc-incVel), and (*3*) to calculate VDGV (acc-VDGV).

IMU-derived measurements were obtained using a Kalman Filter with gyroscope bias estimation to combine raw accelerometer and gyroscope data. Details of the algorithm and its tuning coefficients can be found elsewhere (Chen et al., 2020). The Kalman Filter output provides the direction of gravity in the sensor frame theoretically unaffected by increased motion speeds. Similar to omc-incVel and omc-VDGV, 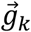 is obtained from IMU was obtained using (*19*) in Appendix C before using (*1*) and (*2*) to calculate incVel (imu-incVel) and (*3*) to calculate VDGV (imu-VDGV). The GVM was calculated directly from unfiltered angular velocity data from the IMU’s gyroscope. This measurement will be referred to as imu-GVM.

#### Angular Velocity Accuracy Assessment

Consistent with our previous studies (Chen et al., 2020, 2018), the offset between the local OMC coordinate frame and the local IMU coordinate frame was determined using angular velocity measurements according to (de Vries et al., 2009). Peak error was defined as the 99^th^ percentile measurement of the rectified sample-to-sample difference between the reference (OMC) and sensor-derived measurements. RMS error was calculated as follows:

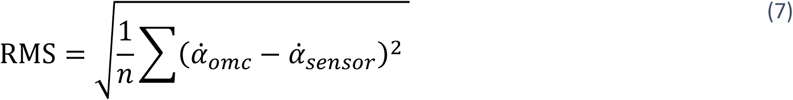

where 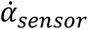 is the sensor-derived angular velocity (i.e., acc-incVel, imu-incVel, acc-VDGV, imu-VDGV, or imu-GVM) and 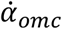 is the corresponding angular velocity calculated from the OMC (i.e., omc-incVel, omc-VDGV, or omc-GVM). Note that the calculation of VDGV measurements rectifies angular velocity.

## Results

As expected, greater angular velocities were observed using omc-GVM compared to omc-VDGV and omc-incVel. Across participants and transfer rates, the mean omc-VDGV was 89% of the mean omc-GVM, and the mean omc-incVel was 77% of the mean omc-GVM. The same trend (i.e., GVM > VDGV > incVel) was observed for the IMU- and accelerometer-based approaches averaged across all transfer rates for the 10th, 50th, 90th, and 99th percentiles of the measured angular velocity values (Figure 1-4). The angular velocities calculated using omc-GVM, omc-VDGV, and omc-incVel from five seconds of a single trial with a ‘fast’ transfer rate are shown in Figure 5.

**Figure 1:**
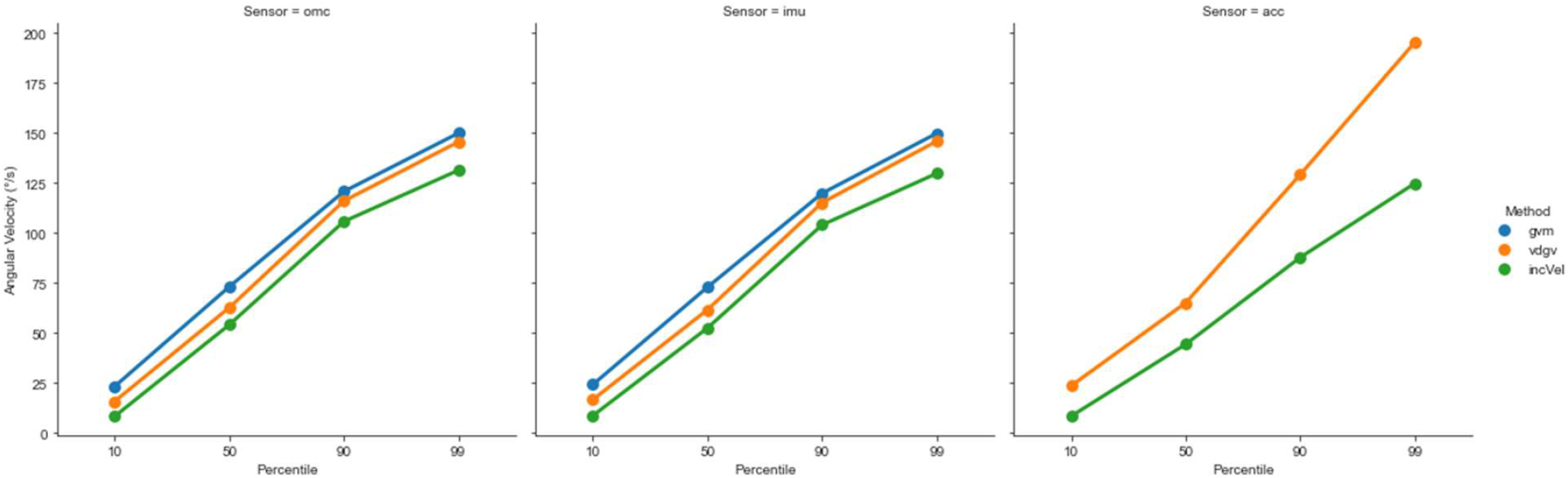
Mean 10^th^, 50^th^, 90^th^, and 99^th^ percentile angular velocities for each sensing modality (omc, imu, and acc) and angular velocity calculation method (gvm, vdgv, and incVel) across all transfer rates.

**Figure 2:**
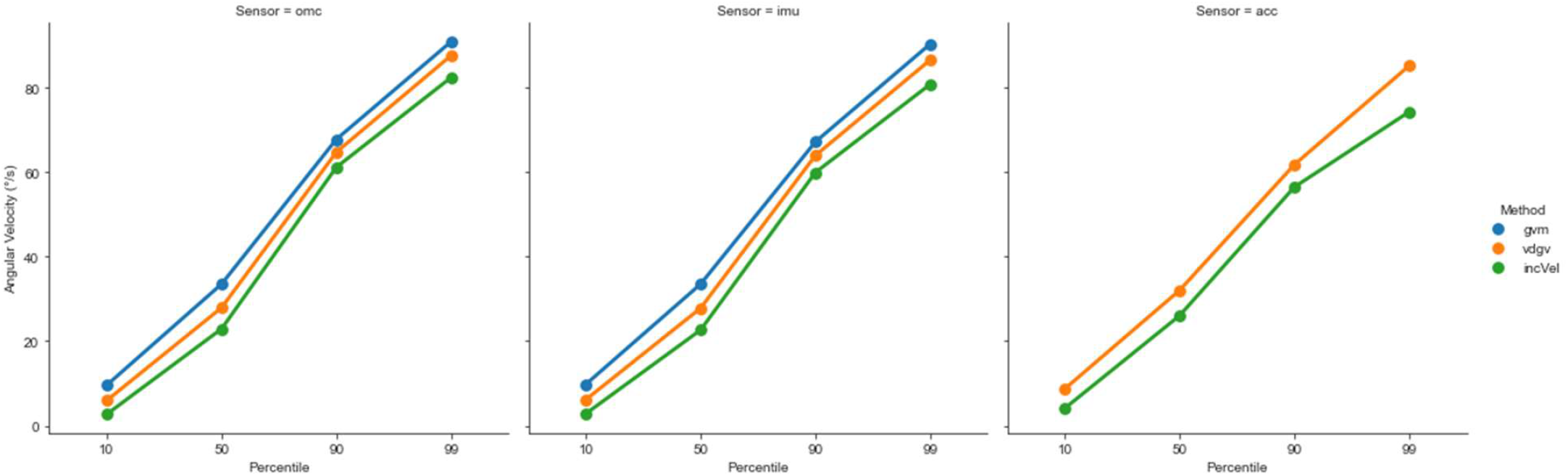
Mean 10^th^, 50^th^, 90^th^, and 99^th^ percentile angular velocities for each sensing modality (omc, imu, and acc) and angular velocity calculation method (gvm, vdgv, and incVel) for the ‘slow’ transfer rate.

**Figure 3:**
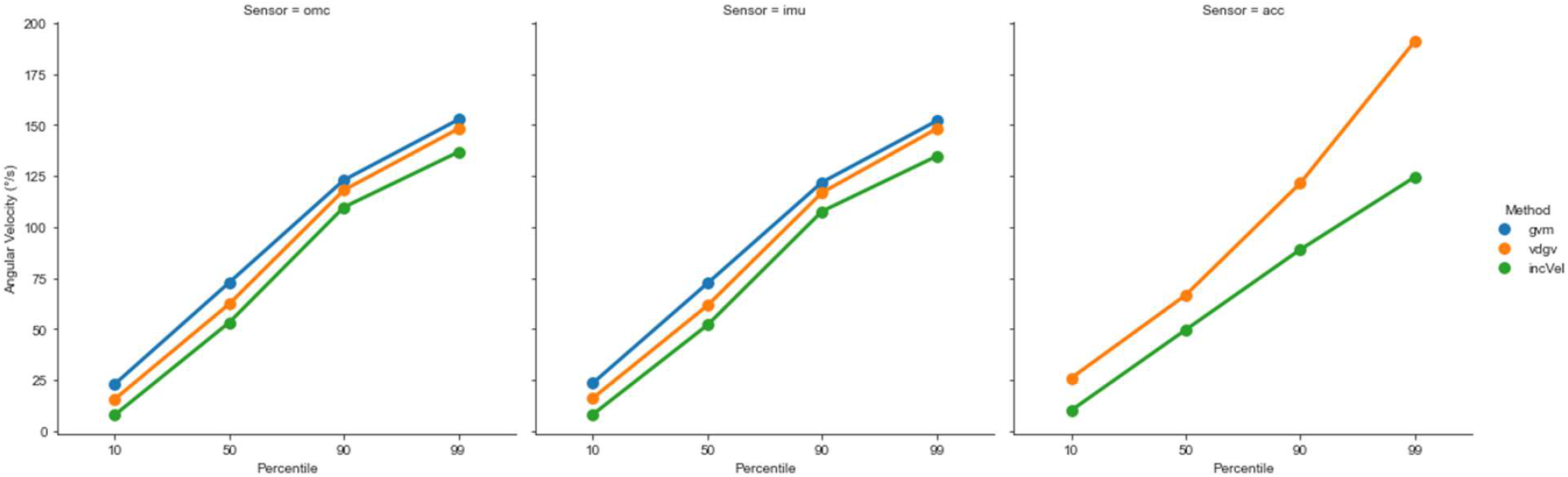
Mean 10^th^, 50^th^, 90^th^, and 99^th^ percentile angular velocities for each sensing modality (omc, imu, and acc) and angular velocity calculation method (gvm, vdgv, and incVel) for the ‘medium’ transfer rate.

**Figure 4:**
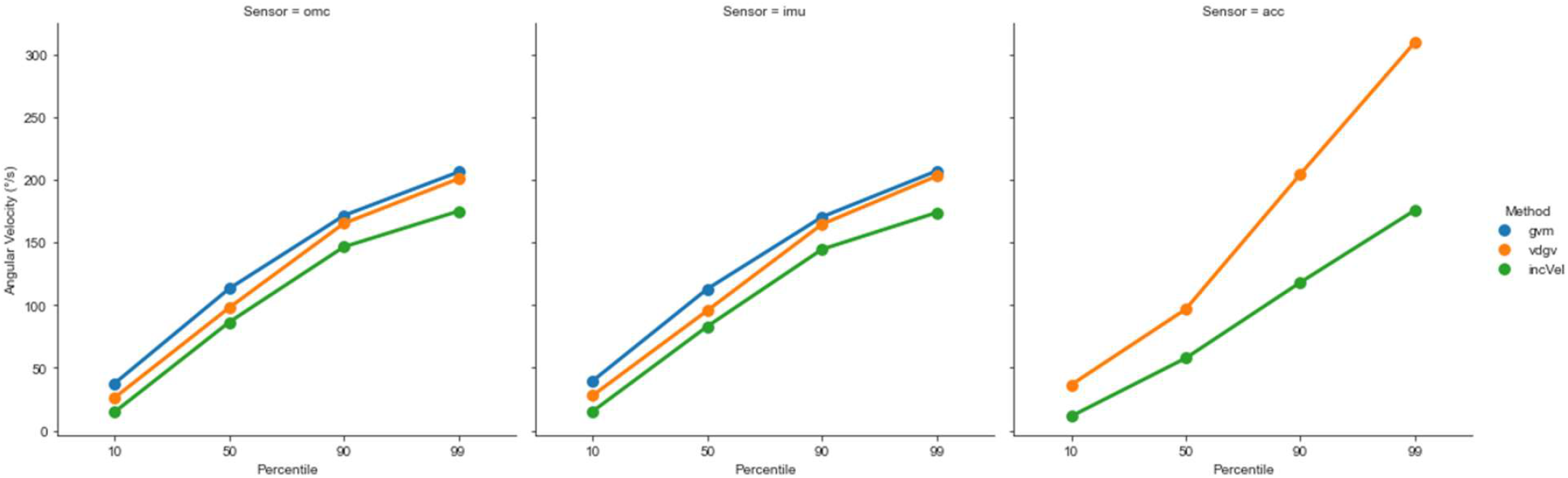
Mean 10^th^, 50^th^, 90^th^, and 99^th^ percentile angular velocities for each sensing modality (omc, imu, and acc) and angular velocity calculation method (gvm, vdgv, and incVel) for the ‘fast’ transfer rate.

**Figure 5:**
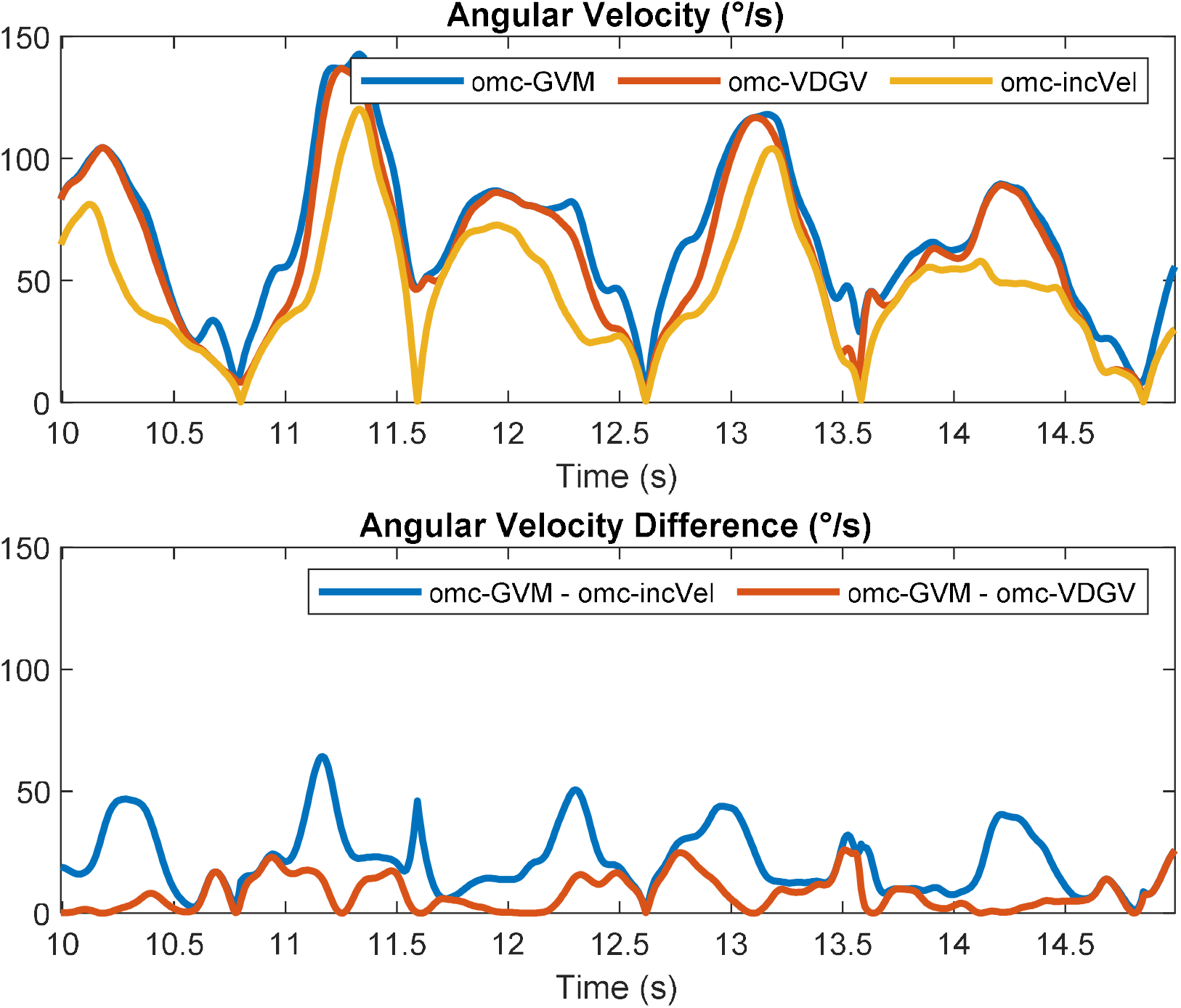
[top] Optical motion capture-derived angular velocities from five seconds of a single trial with ‘fast’ transfer rate calculated using gyro vector magnitude (omc-GVM), vector-differenced generalized velocity (omc-VDGV), and inclination velocity (omc-incVel). [bottom]: Sample-to-sample difference between omc-GVM and omc-incVel, as well as omc-GVM and omc-VDGV.

**Figure 6:**
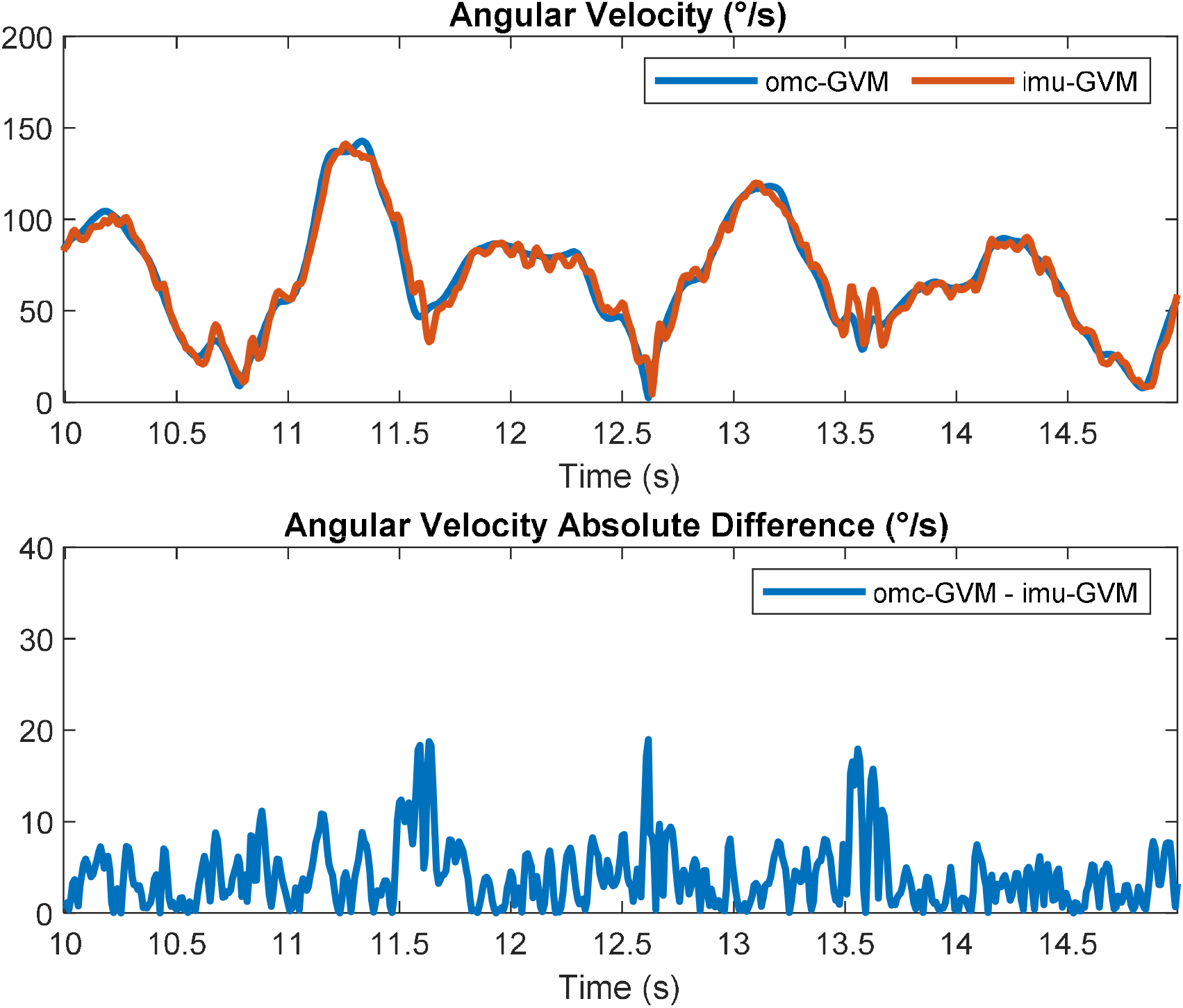
[top] Angular velocity from five seconds of a single trial with ‘fast’ transfer rate calculated using gyro vector magnitude calculated from optical motion capture measurements (omc-GVM) and from a gyroscope contained within the inertial measurement unit (imu-GVM). [bottom] Sample-to-sample difference between omc-GVM and imu-GVM.

**Figure 7:**
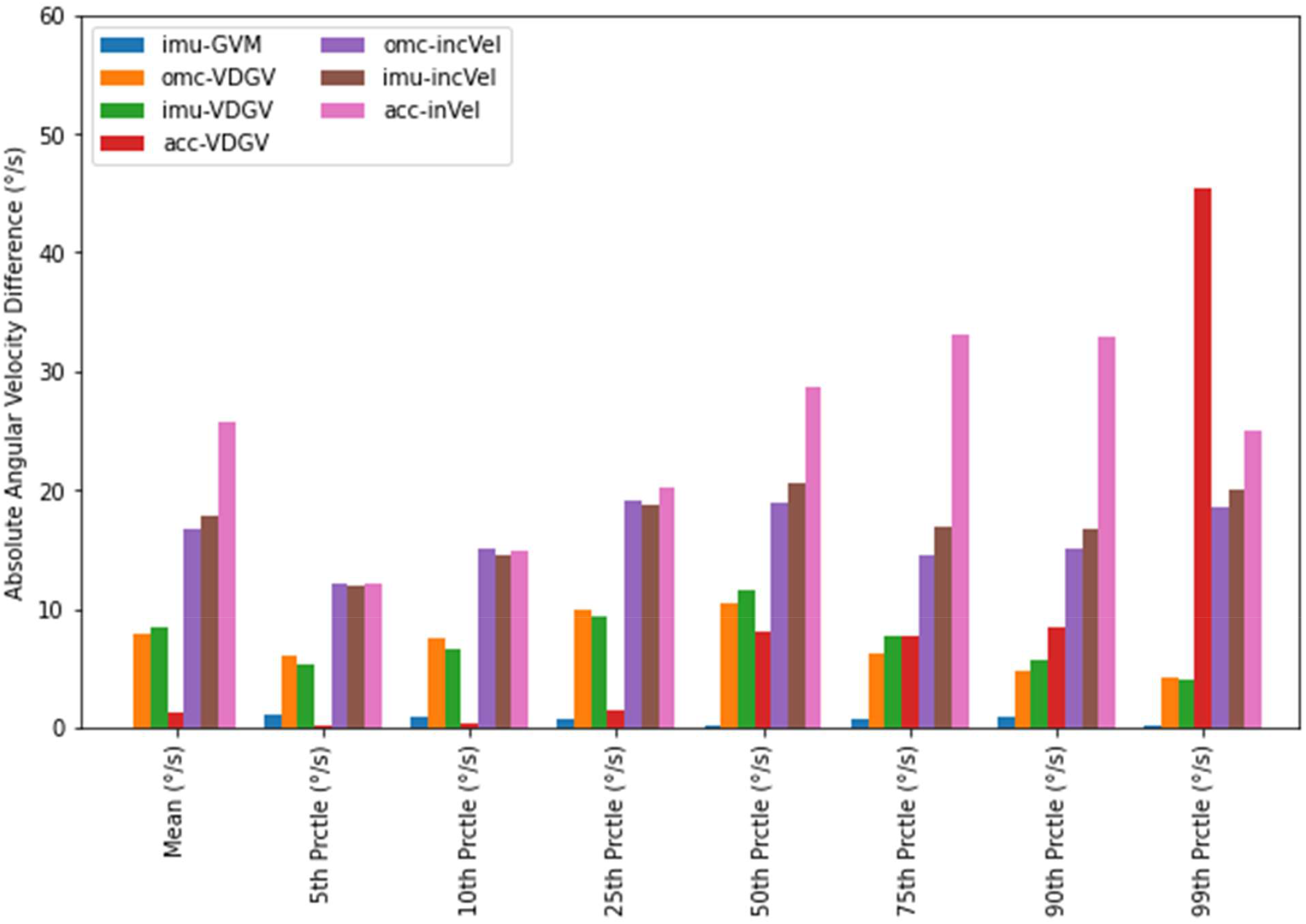
Absolute angular velocity difference between optical motion capture-derived gyroscope vector magnitude and various angular velocity calculation method across all transfer rates.

**Figure 8:**
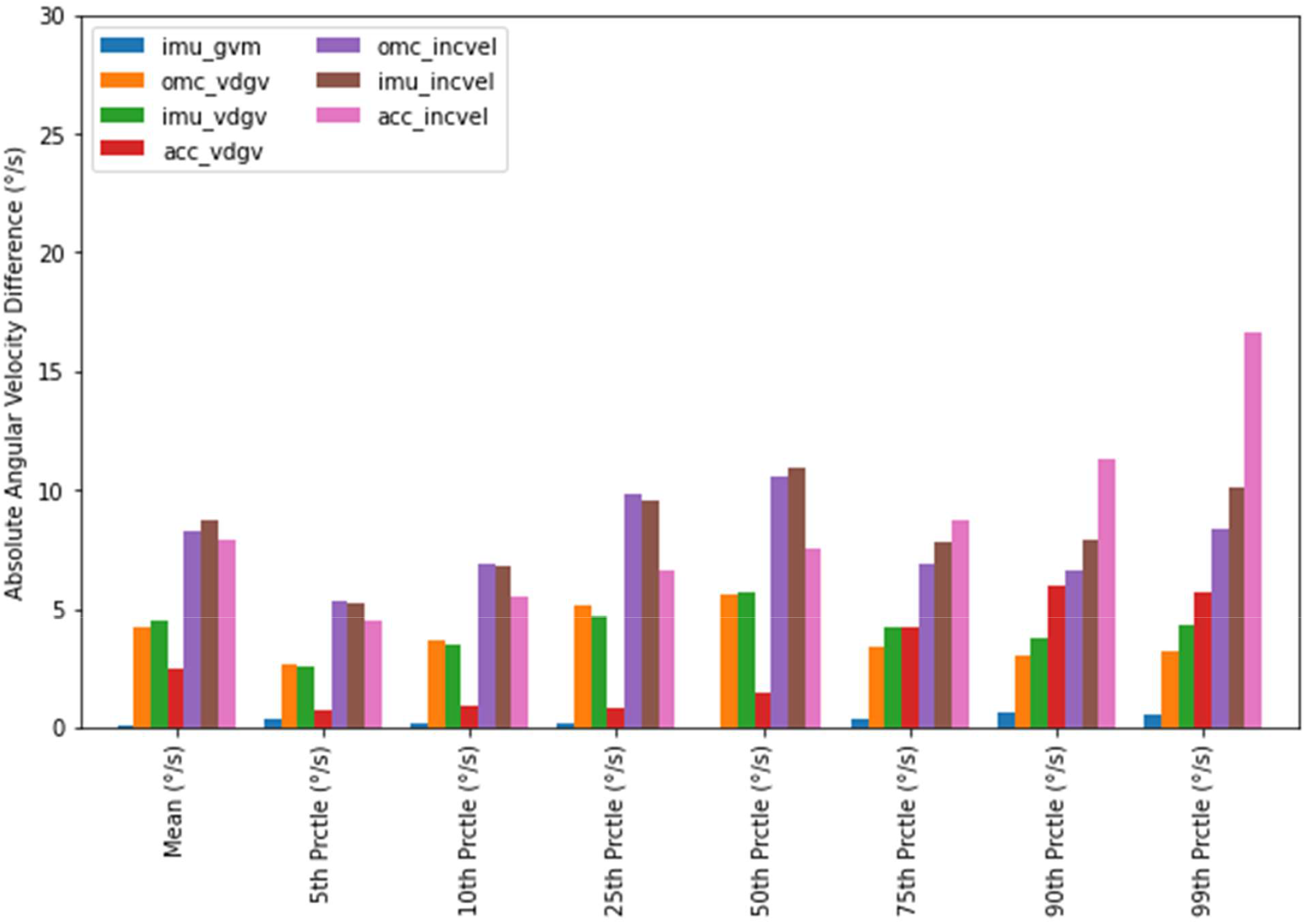
Absolute angular velocity difference between optical motion capture-derived gyroscope vector magnitude and various angular velocity calculation method for ‘slow’ transfer rate.

**Figure 9:**
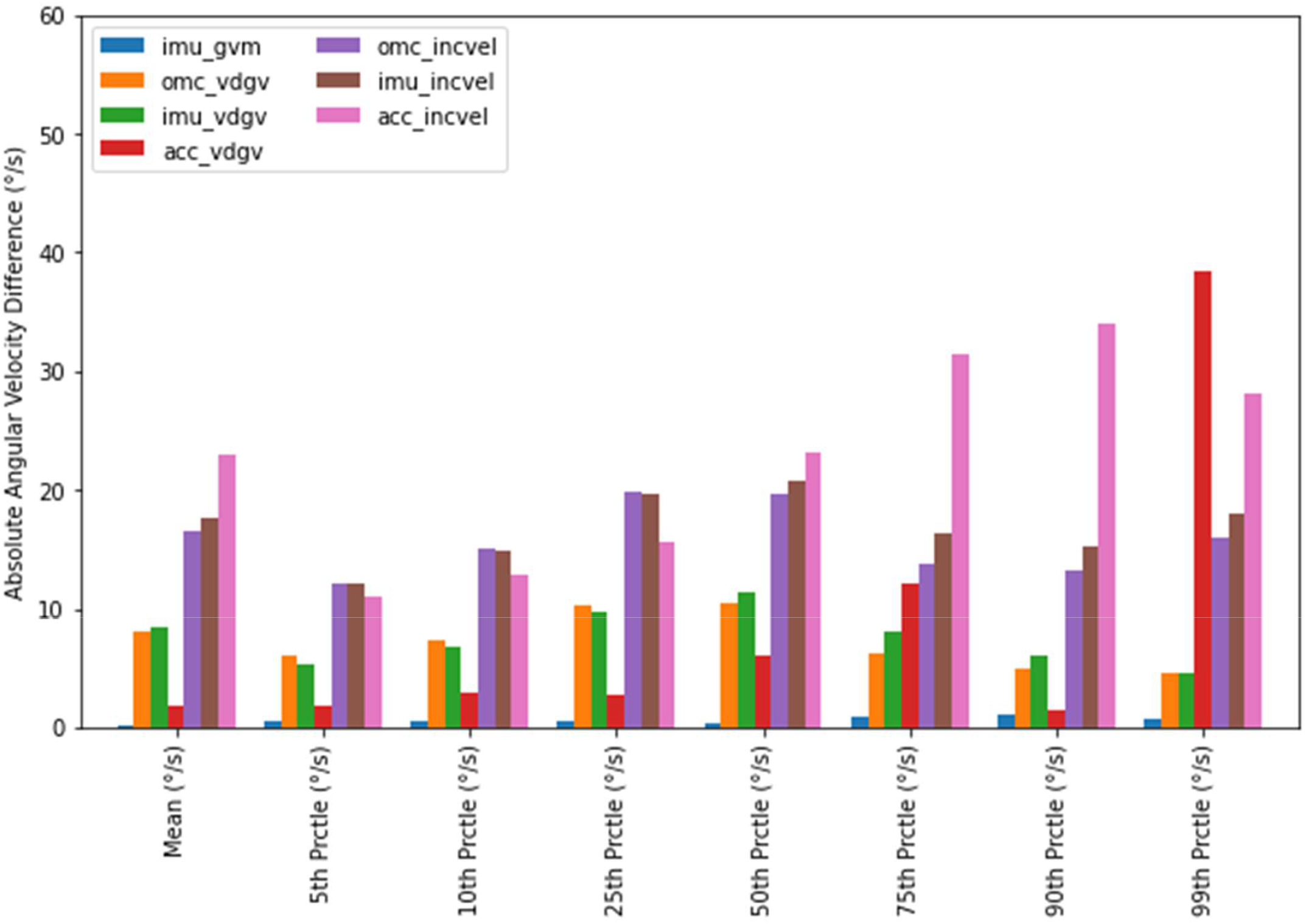
Absolute angular velocity difference between optical motion capture-derived gyroscope vector magnitude and various angular velocity calculation method for ‘medium’ transfer rate.

**Figure 10:**
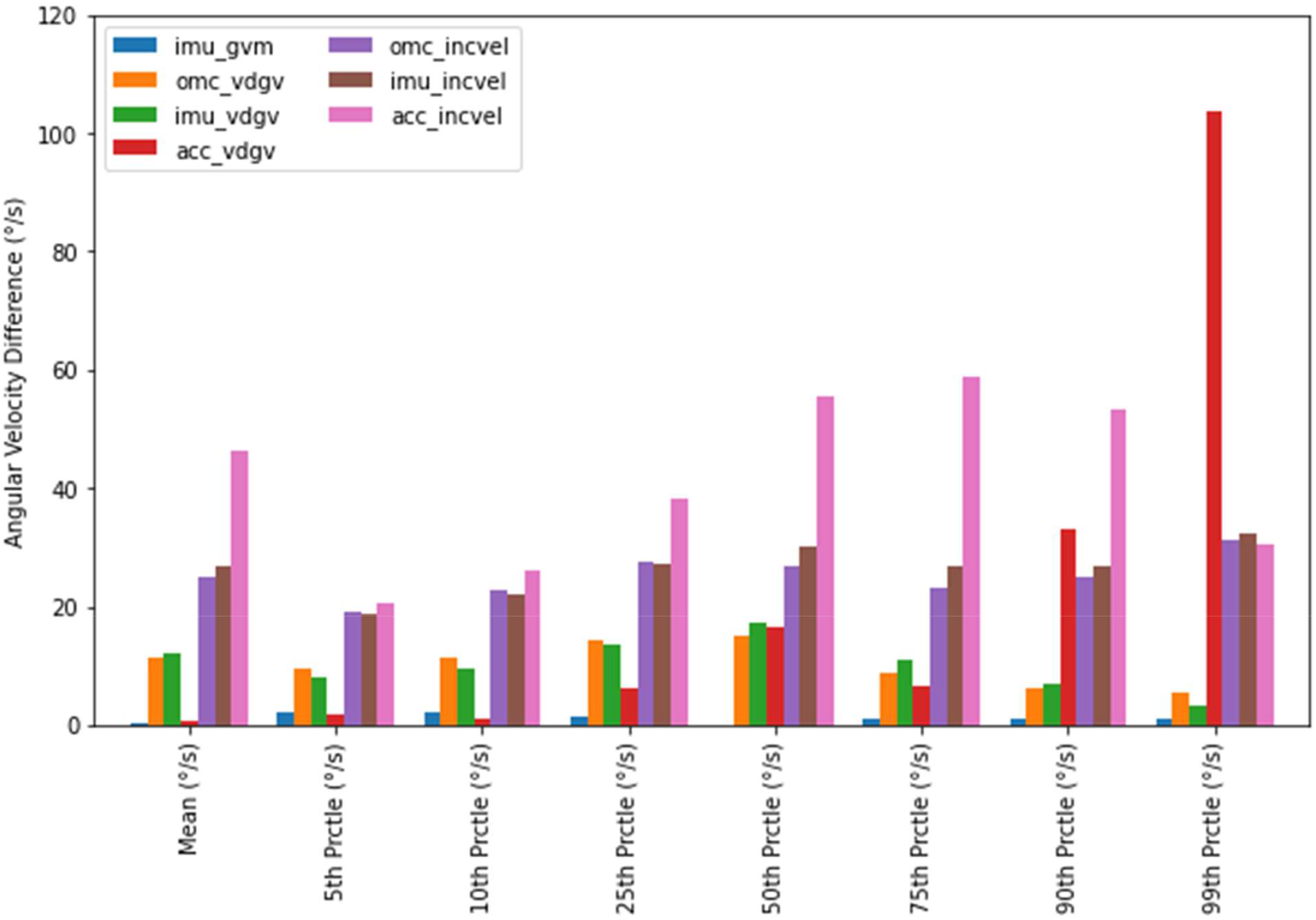
Absolute angular velocity difference between optical motion capture-derived gyroscope vector magnitude and various angular velocity calculation method for ‘fast’ transfer rate.

**Figure 11:**
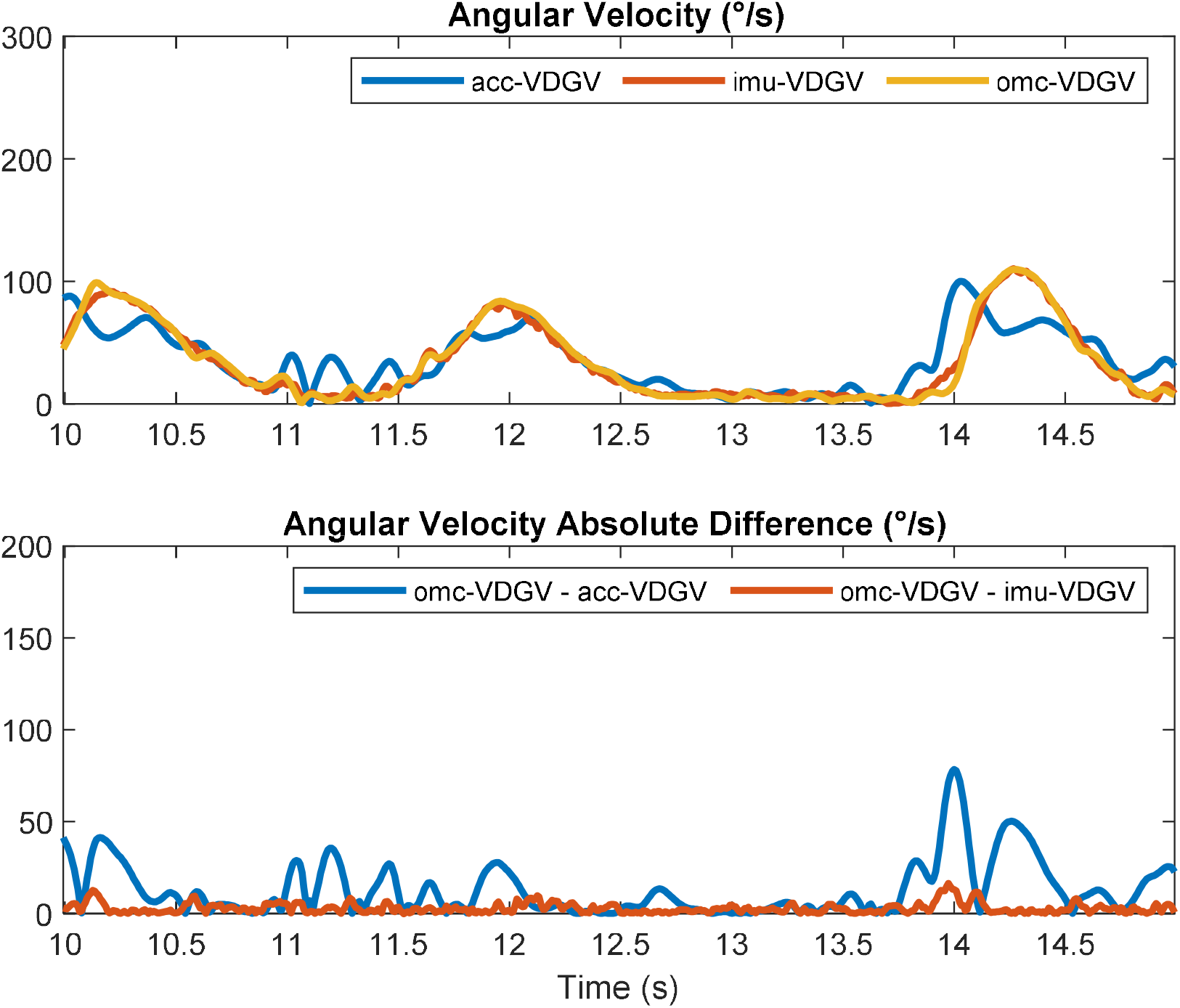
[top] Vector-differenced generalized velocities measured from an optical motion capture system (omc-VDGV), inertial measurement unit (imu-VDGV), and accelerometer (acc-VDGV) from five seconds of a single trial with ‘slow’ transfer rate. [bottom]. Sample-to-sample absolute difference between omc-VDGV and acc-VDGV, and omc-VDGV and imu-VDGV.

**Figure 12:**
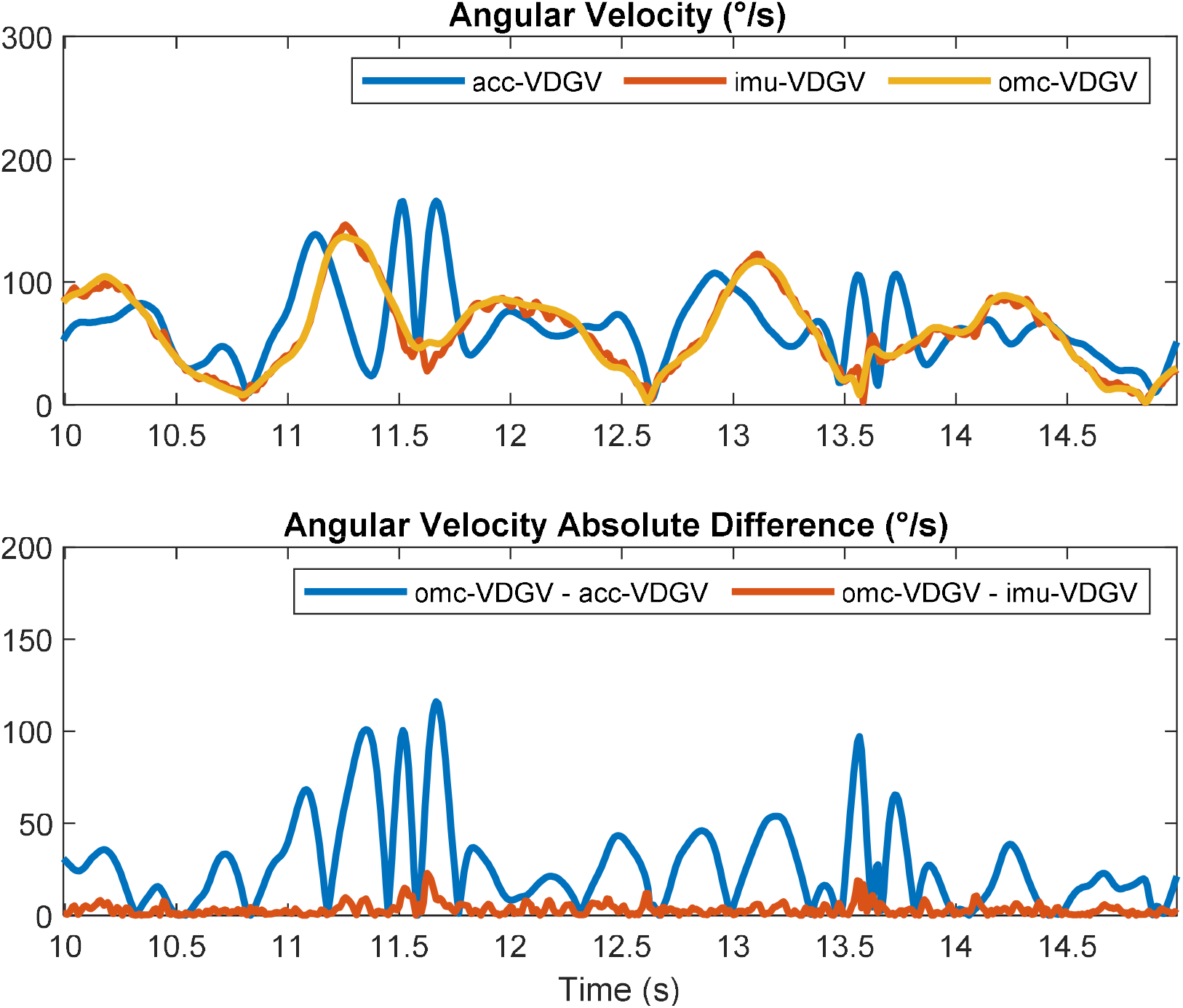
[top] Vector-differenced generalized velocities measured from an optical motion capture system (omc-VDGV), inertial measurement unit (imu-VDGV), and accelerometer (acc-VDGV) from a single trial with ‘fast’ transfer rate. [bottom]. Sample-to-sample absolute difference between omc-VDGV and acc-VDGV, and omc-VDGV and imu-VDGV.

Mean RMS and peak angular velocity errors by transfer rate, angular velocity calculation method, sensing modality, and reference signal are shown in Table 1. Using omc-GVM as the reference, RMS and peak errors for omc-VDGV were about 18°/s and 56°/s, respectively, for the ‘fast’ transfer rate. RMS and peak errors for omc-incVel were about 32°/s and 89°/s, respectively. These results provide empirical support for the notion that, in the absence of error introduced through the use of accelerometer data, VDGV yields lower measurement error than incVel.

**Table 1:** Mean(SD) root-mean-square and peak error angular velocities across 13 participants and various transfer rates calculated using three different methods: gyro vector magnitude calculated using optical motion capture (omc-GVM), inertial measurement unit (imu-GVM); vector-differenced generalized velocity (VDGV) using optical motion capture (omc-VDGV), inertial measurement unit (imu-VDGV), accelerometer (acc-VDGV); inclination velocity calculated using optical motion capture (omc-incVel), inertial measurement unit (imu-incVel), and accelerometer (acc-incVel).

| IMU-incVel | Reference Computational Method |  |  |  |  |  |
| --- | --- | --- | --- | --- | --- | --- |
|  | <u>omc-GVM</u> |  | <u>omc-VDGV</u> |  | <u>omc-incVel</u> |  |
|  | <u>RMS error</u> | <u>Peak error</u> | <u>RMS error</u> | <u>Peak error</u> | <u>RMS error</u> | <u>Peak error</u> |
|  | (°/s) | (°/s) | (°/s) | (°/s) | (°/s) | (°/s) |
|  | mean (sd) | mean (sd) | mean (sd) | mean (sd) | mean (sd) | mean (sd) |
| **‘Slow’ Rate** |  |  |  |  |  |  |
| omc-VDGV | 7.0(1.8) | 25.0(6.5) | -REF- | -REF- | 7.3(2.9) | 29.2(12.5) |
| omc-incVel | 12.0(3.2) | 40.8(12.8) | 7.3(2.9) | 29.2(12.5) | -REF- | -REF- |
| imu-GVM | 3.5(0.7) | 11.4(2.6) | 7.8(1.9) | 26.2(6.9) | 12.7(3.2) | 43.6(13.3) |
| imu-VDGV | 7.7(1.8) | 25.5(6.3) | 3.5(1.1) | 11.9(4.1) | 8.2(2.8) | 31.8(12.3) |
| acc-VDGV | 12.6(2.6) | 41.8(11.7) | 13.0(3.1) | 44.9(13.8) | 15.7(3.8) | 56.3(18.2) |
| imu-incVel | 12.4(3.2) | 40.5(12.5) | 8.0(2.7) | 29.9(12.0) | 3.0(1.0) | 9.6(3.8) |
| acc-incVel | 15.3(3.1) | 47.5(12.6) | 13.0(3.0) | 41.7(11.9) | 11.7(2.6) | 38.4(10) |
| **‘Medium’ Rate** |  |  |  |  |  |  |
| omc-VDGV | 12.9(3.1) | 42.4(10.4) | -REF- | -REF- | 14.3(5.1) | 53.0(21.8) |
| omc-incVel | 22.6(6.1) | 67.7(21.1) | 14.3(5.1) | 53.0(21.8) | -REF- | -REF- |
| imu-GVM | 5.7(1.4) | 18.8(5.8) | 14.4(3.2) | 46.5(11) | 24.1(6.5) | 77.0(24.7) |
| imu-VDGV | 14.0(2.8) | 43.7(9.8) | 6.5(1.6) | 22.2(6.5) | 15.7(5.0) | 58.5(21.4) |
| acc-VDGV | 39.8(10.5) | 118.8(38.5) | 44.0(11.6) | 135.6(44.3) | 49.0(13.9) | 154.0(51.5) |
| imu-incVel | 23.6(5.7) | 68.1(19.9) | 15.4(4.6) | 52.7(19.1) | 5.4(1.4) | 16.0(4.5) |
| acc-incVel | 39.8(8.0) | 106.2(18.1) | 36.3(8.7) | 99.5(19.7) | 33.4(9.0) | 94.4(22.6) |
| **‘Fast’ Rate** |  |  |  |  |  |  |
| omc-VDGV | 17.7(8.1) | 55.6(23.4) | -REF- | -REF- | 21.2(7.6) | 71.0(25.4) |
| omc-incVel | 32.3(11.4) | 89.4(30.7) | 21.2(7.6) | 71.0 (25.4) | -REF- | -REF- |
| imu-GVM | 8.3(1.7) | 27.9(6.9) | 20.7(7.4) | 64.8(21.1) | 34.9(11.3) | 103.7(32.3) |
| imu-VDGV | 20.1(7.2) | 59.5(22) | 9.9(2.3) | 32.9(9.4) | 23.7(7.2) | 81.4(25.0) |
| acc-VDGV | 78.9(15.9) | 210.3(59.3) | 85.6(19) | 235.9(69.2) | 90.0 (23.1) | 257.0 (79.4) |
| imu-incVel | 34.2(10.6) | 90.9(28) | 23.3(6.9) | 72.0 (22.2) | 8.0 (2) | 23.1(6.9) |
| acc-incVel | 72.0 (11.0) | 164.6(23.2) | 66.7(9.2) | 159.5(22) | 59.8(9.3) | 146.1(19.5) |

Theoretically, then, and assuming omc-VDGV is the “true” VDGV (i.e., as the reference signal), one would expect to observe lower error for acc-VDGV than for acc-incVel. However, acc-VDGV and acc-incVel exhibited similar RMS and peak errors at the ‘slow’ transfer rate and, at both the ‘medium’ and ‘fast’ transfer rates, both RMS and peak errors were lower for acc-incVel than for acc-VDGV (e.g., at the ‘fast’ transfer rate, peak error was about 160°/s for acc-incVel but about 236°/s for acc-VDGV). As expected, incorporating the gyroscope and sensor fusion dramatically reduced error relative to the omc-VDGV reference. For example, at the ‘fast’ transfer rate, the peak error for imu-VDGV was about 33°/s compared to 236°/s for acc-VDGV.

Assuming omc-GVM reflects the true “total” angular velocity of the upper arm, imu-GVM yielded the lowest error among all sensor-based approaches for angular velocity measurement considered in this study. Error magnitudes for imu-GVM were also the least susceptible to influence from motion speed. The mean RMS error of imu-GVM increased from 3.5°/s at the ‘slow’ transfer rate to 8.3°/s at the ‘fast’ transfer rate (a 2.4-fold increase). In contrast, the mean RMS error of imu-VDGV increased from 7.7°/s to 20.1°/s (a 2.6-fold increase), the mean RMS error of imu-incVel increased from 12.4°/s to 34.2°/s (a 2.8-fold increase), the mean RMS error of acc-incVel increased from 15.3°/s to 72.0°/s (a 4.7-fold increase), and the mean RMS error of acc-VDGV increased from 12.6°/s to 78.9°/s (a 6.3-fold increase). The mean peak error was also the lowest for imu-GVM across all transfer rates, with an increase from 11.4°/s at the ‘slow’ transfer rate to 27.9°/s at the ‘fast’ transfer rate (a 2.6-fold increase). The mean peak errors for imu-VDGV and imu-incVel were greater than for imu-GVM. However, the proportional increases observed were similar (i.e., 2.3-fold for imu-VDGV and 2.2-fold for imu-incVel). In addition to greater mean peak error than imu-GVM, the proportional increases for acc-VDGV and acc-incVel were more dramatic (i.e., 5.0-fold for acc-VDGV and 3.5-fold for acc-incVel).

## Discussion

Both acc-incVel and acc-VDGV were conceived as methods to measure the speed of body segments in occupational health studies when miniature, inexpensive gyroscopes were not prevalent in wearable devices. However, acc-incVel and acc-VDGV have two inherent limitations: (i) the angular velocities captured do not reflect the full extent of motion (confirmed in this study with mean omc-incVel and omc-VDGV velocities that are 77% and 89% of the mean omc-GVM velocities), and (ii) the accuracy of accelerometer-based angular velocity measurements is reduced as motion speeds increase (indicated in this study as a ∼108°/s increase in peak error of acc-incVel [vs. omc-incVel) and a ∼190°/s increase in peak error of acc-VDGV [vs. omc-VDGV] from the slow to fast transfer rates). Using GVM addresses both limitations, as indicated by (i) peak errors of <30°/s in imu-GVM [vs. omc-GVM] across all motion speeds, and (ii) just a ∼16°/s increase in peak error from the slow to fast transfer rates.

Although all three accelerometer axes are used in the calculations of both incVel and VDGV, neither method captures all three directions of motion. For example, pure rotation around gravity will yield zero angular velocity when calculated using either VDGV or incVel. VDGV may be preferable to incVel since it captures a fuller extent of motion. However, the results of the current study suggest some important methodological limitations. We did not expect greater error for acc-VDGV compared to acc-incVel when considering omc-VDGV as the reference (for the ‘medium’ and ‘fast’ transfer rates). We believe the most likely explanation for this result is a ‘compounding’ of accelerometer error across multiple movement directions (i.e., two movement directions with VDGV vs. one with incVel) as motion speeds increase. Although the inclusion of a gyroscope and sensor fusion (i.e., imu-VDGV and imu-incVel) resulted in superior error characteristics, these approaches can still not capture rotation around gravity.

The results of this study verify that angular velocity measurements calculated using GVM encompass all movement directions and can be easily calculated using measurements from field-capable IMUs. When omc-GVM was used as the reference, imu-GVM produced the most accurate sensor-based angular velocity measurements, with peak errors <27.9°/s across all transfer rates. This is in contrast to acc-VDGV measurements, which theoretically better represents the “true” angular velocity compared to incVel but may not empirically as indicated in this study. Errors associated with angular velocities measured from gyroscopes are time-invariant and that gyroscopic drift is introduced when gyroscope measurements are integrated with respect to time to obtain spatial orientation without the use of a sensor fusion algorithm. Although omc-GVM was used as the reference, we hypothesize that the slight increase in imu-GVM errors with increased transfer rates may be attributed to the differentiation of OMC-derived orientation measurements with respect to time, which itself introduces error that may be magnified as the transfer rate increased. It is conceivable that the OMC can produce more accurate orientation measurements while the gyroscope produces more accurate angular velocities.

The relative magnitude between GVM, VDGV, and incVel measurements is generally difficult to compare across studies due to differences in motion patterns. GVM, VDGV, and incVel will produce identical results with planar motion in the direction of incVel. However, only GVM will register angular velocities when motion occurs purely around gravity. To our knowledge, Fan et al. (2021) provided the only comparable study and reported that the mean upper arm imu-VDGV was 16.5°/s greater than the mean upper arm imu-IncVel during manual material handling activities. In our study, the mean omc-VDGV was 4.2°/s, 8.7°/s, and 13.8°/s greater than the mean omc-incVel for the ‘slow,’ ‘medium,’ and ‘fast’ transfer rates, respectively. We hypothesize that these differences may be attributed to differences in motion patterns. In our study, a short-duration, cyclic task was performed in contrast to actual material handling tasks, which are likely to involve greater motion complexity. Furthermore, Fan et al. (Fan et al., 2021) calculated VDGV and IncVel using sensor fusion from IMU measurements. Although sensor fusion substantially reduces the effects of increased motion speeds (i.e, increased non-gravitational acceleration) on angular velocity accuracy, the presence of non-gravitational acceleration will adversely affect the accuracy of angular velocities derived from vector measurements since accelerometer measurements are used. Fan et al. (2021) reported acc-VDGV mean and percentile measurements generally greater than imu-VDGV, with the 90^th^ and 99^th^ percentile acc-VDGV exceeding imu-VDGV by over 100°/s and 200°/s, respectively. The results were consistent with our study, which indicated that acc-VDGV mean and percentile measurements were generally greater than imu-VDGV, with observed percentile differences >100°/s. However, in our study, the difference between mean imu-VDGV and acc-VDGV was <15°/s across all transfer rates, in contrast to the 50°/s difference observed in Fan et al. (2021) despite a mean imu-VDGV that was lower than our ‘medium’ transfer rate. We again hypothesize that our observed differences between VDGV and incVel are smaller since our motion pattern was less complex.

### Strengths and Limitations

An important strength of the current study is our use of OMC measurements for comparing GVM, VDGV, and incVel, which provided ‘true’ reference signals for comparison (Cuesta-Vargas et al., 2010) and controlled for potential IMU errors when evaluating angular velocity calculation methods. The comparisons of acc-VDGV and imu-VDGV to omc-VDGV also highlight potential inaccuracies of acc-VDGV and the improvements that can be achieved through the use of sensor fusion (e.g. (Chen et al., 2020, 2018)). Furthermore, the derivations provided a geometric explanation for a quantity of angular velocity sometimes found in the literature (i.e., GVM) and how it relates to the generalized velocity calculations often used to estimate movement velocities in studies of occupational health. The main limitation of this paper is the relatively simplistic motion pattern. We expect differences between GVM, VDGV, and incVel to be accentuated as motion complexity increases. Furthermore, the error magnitudes observed may not generalize to a field-based study (i.e., in a workplace) since the motion speeds tested are unlikely to be sustained beyond a few minutes.

### Methodological Considerations for Future Studies

When considering historical data captured with accelerometers, the difference between acc-incVel and acc-VDGV may not be appreciable, particularly at high movement speeds. In these cases, acc-incVel will likely produce angular velocities more representative of ‘true’ VDGV than acc-VDGV, given the sensitivity of acc-VDGV to non-gravitational acceleration. Furthermore, the implication that acc-VDGV tends to over-estimate velocities compared to omc-VDGV, particularly near the extremes of the angular velocity distribution (e.g., the 90^th^ and 99^th^ percentiles), could contribute to error in the estimates of the dose-response relationship between exposures to high angular velocities and WMSDs, such as those provided by Arvidsson et al. (2021). The differences in imu-VDGV and acc-VDGV may also result in difficulties when comparing angular velocities across historical studies. Angular velocities calculated using imu-VDGV may, for example, be considered ‘safe’ according to proposed threshold limit values while considered ‘exposed’ when derived from acc-VDGV.

Given the availability of gyroscope measurements, we suggest GVM as the preferred angular velocity calculation method in future studies since it captures all three motion directions and is theoretically unaffected by either increased motion speeds or measurement duration. We also suggest quantifying imu-VDGV, accel-VDGV, imu-incVel, and acc-incVel to facilitate comparison across historical studies as technology continues to improve. The additional information provided in Appendix D can aid this process, particularly refining conversion models that have been previously proposed (Forsman et al., 2022b).

## Data Availability

All data produced in the present work are contained in the manuscript

## Acknowledgments

This study was supported by research funding from the Centers for Disease Control (CDC)/National Institute for Occupational Safety and Health (NIOSH). This included funding from the Heartland Center for Occupational Health and Safety at the University of Iowa (T42OH008491), the Deep South Center for Occupational Health and Safety at the University of Alabama-Birmingham (UAB) and Auburn University (T42OH008436). The findings and conclusions in this report are those of the authors and do not necessarily represent the views of the CDC/NIOSH.

## APPENDIX A Quaternion Basics

Rotating a 3-dimensional vector from frame 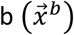 to frame 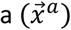 is accomplished as follows:

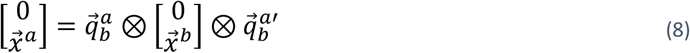

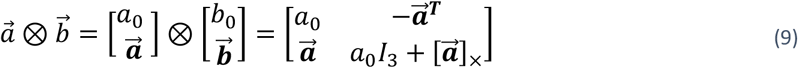

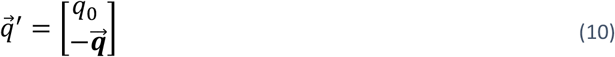

where ⊗ is the quaternion product, 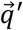 is the quaternion conjugate of 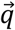, [·]_×_is the skew symmetric operator, and *I*_3_ is a 3×3 identity matrix.

Similarly, rotating a 3-dimensional vector from frame a to frame b is achieved as follows:

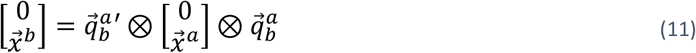

Given 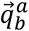 which describes the rotation from frame b to frame a, and 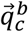 which describes the rotation from frame c to frame b, 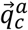, which describes the rotation from frame c to frame a, is calculated as follows:

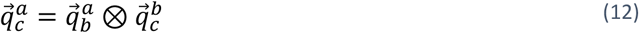

Similarly, given 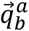 and 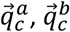, can be obtained as follows:

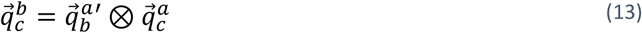

## Appendix B Defining Gyroscope Vector Magnitude

From Equations (*12*) and (*13*), 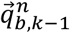 and 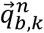 can be defined as the orientation at instance k-1 and k relative to a common reference frame *n*, and 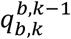 can be defined as the orientation of 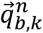 relative to 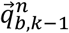. Given an initial orientation and perfect angular velocities in the sensor frame, angular velocities can be integrated with respect to time by applying Equation (*14*) recursively to calculate subsequent orientation (Brodie et al., 2008).

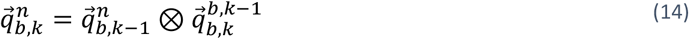

Since 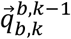 describes the orientation, it can be decomposed into a direction specified by the unit vector 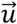 and angle η (*15*). When 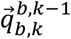 is determined using angular velocity measurements in the sensor frame (i.e., gyroscope measurements), 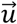 is calculated by normalizing 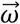 to unitary (*16*) and *η* is determined by multiplying the vector magnitude of 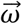 by its sample period (Δ*t*), as seen in Equation (*18*). *η* can be therefore be defined as the net directional change between successive measurements of orientation.

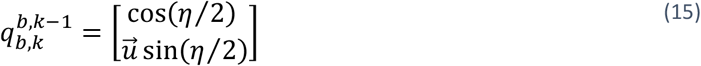

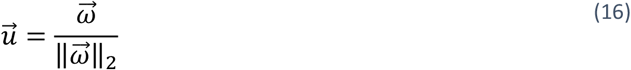

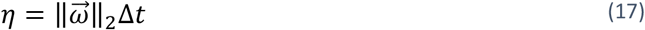

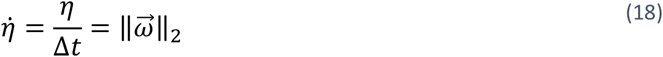

Dividing *η* by its sample period would yield the rate of directional change between successive measurements of orientation 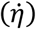, also known as the angular velocity. Therefore, the vector magnitude of gyroscope measurements can be defined as the rate of directional change between successive orientation measurements.

## Appendix C Calculating gravitational vector and angular velocities from a quaternion

An OMC or the sensor fusion output from an IMU will provide quaternion 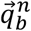 which describes the spatial orientation of a given rigid body (*b*) relative a reference frame (*n*). Given 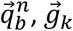, can be calculated by applying (*11*) as follows:

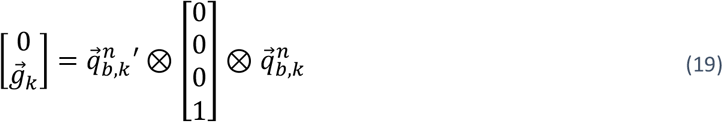

Given 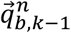 and 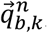, and Δ*t*, 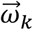 can be calculated as follows:

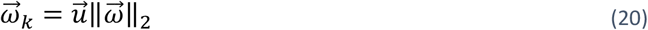

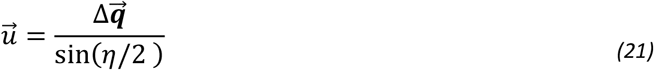

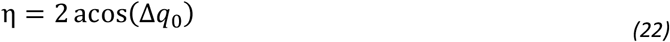

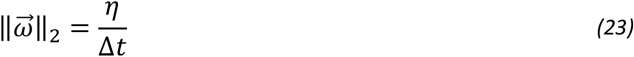

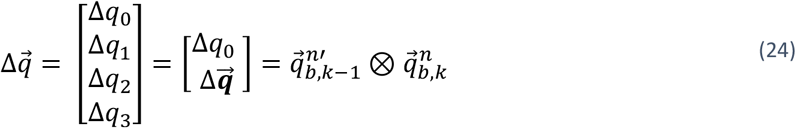

## Appendix D Angular velocity summary metrics

**Table 2:** Mean(SD) angular velocities across 13 participants and various transfer rates calculated using various methods: gyro vector magnitude calculated using optical motion capture (omc-GVM), inertial measurement unit (imu-GVM); vector-differenced generalized velocity (VDGV) using optical motion capture (omc-VDGV), inertial measurement unit (imu-VDGV), accelerometer (acc-VDGV); inclination velocity calculated using optical motion capture (omc-incVel), inertial measurement unit (imu-incVel), and accelerometer (acc-incVel).

|  | omc-GVM | imu-GVM | omc-VDGV | imu-VDGV | acc-VDGV | omc-incVel | imu-incVel | acc-incVel |
| --- | --- | --- | --- | --- | --- | --- | --- | --- |
| **‘Slow’ Transfer Rate** |  |  |  |  |  |  |  |  |
| Mean (°/s) | 36.6(2.9) | 36.5(2.9) | 32.2(2.7) | 32.0(2.6) | 34.0(3.1) | 28.2(2.1) | 27.7(2.0) | 28.6(2.3) |
| 5th Percentile (°/s) | 6.5(2.5) | 6.8(2.7) | 3.7(1.6) | 3.9(1.7) | 5.8(1.7) | 1.3(0.6) | 1.3(0.6) | 2.1(0.7) |
| 10th Percentile (°/s) | 9.5(3.5) | 9.7(3.5) | 5.8(2.3) | 6.0(2.4) | 8.5(2.2) | 2.6(1.1) | 2.7(1.2) | 4.0(1.3) |
| 25th Percentile (°/s) | 17.2(4.7) | 17.5(4.7) | 12.0(3.7) | 12.4(3.6) | 16.3(2.8) | 7.4(2.7) | 7.6(2.6) | 10.6(2.5) |
| 50th Percentile (°/s) | 33.5(4.0) | 33.5(4.0) | 27.8(3.8) | 27.7(3.5) | 31.9(3.4) | 22.8(3.8) | 22.5(3.4) | 25.9(2.7) |
| 75th Percentile (°/s) | 53.3(5.2) | 52.8(5.1) | 49.7(4.8) | 48.9(4.7) | 49.0(5.1) | 46.2(5.0) | 45.2(4.7) | 44.4(4.6) |
| 90th Percentile (°/s) | 68.1(9.0) | 67.4(8.8) | 64.9(8.8) | 64.1(8.7) | 61.9(7.5) | 61.3(8.7) | 60.1(8.4) | 56.6(7.0) |
| 99th Percentile (°/s) | 93.4(16.4) | 92.9(16.5) | 89.9(15.9) | 89.1(16.0) | 86.9(14.7) | 84.4(14.5) | 82.4(14.1) | 75.5(10.7) |
| Time at low velocities (<5°/s)(%) | 4.0(2.7) | 3.7(2.6) | 9.5(4.4) | 9.1(4.3) | 4.6(2.5) | 19.5(6.4) | 19.0(6.2) | 13.2(3.7) |
| Time at high velocities (≥90°/s)(%) | 2.1(2.5) | 2.0(2.4) | 1.6(2.0) | 1.5(1.9) | 1.1(1.4) | 1.0(1.5) | 0.9(1.4) | 0.4(0.6) |
| **‘Medium’ Transfer Rate** |  |  |  |  |  |  |  |  |
| Mean (°/s) | 72.7(6.7) | 72.5(6.8) | 64.6(5.8) | 64.1(5.9) | 70.7(8.1) | 56.0(5.3) | 55.0(5.3) | 49.7(5.9) |
| 5th Percentile (°/s) | 15.8(5.3) | 16.3(5.4) | 9.8(3.8) | 10.4(4.1) | 17.2(5.7) | 3.6(1.5) | 3.8(1.5) | 4.9(1.7) |
| 10th Percentile (°/s) | 22.6(6.5) | 23.2(6.6) | 15.1(5.3) | 15.6(5.7) | 25.1(6.7) | 7.5(2.8) | 7.8(2.8) | 9.8(3.0) |
| 25th Percentile (°/s) | 40.5(8.8) | 41.0(8.8) | 30.1(7.3) | 30.6(7.5) | 43.2(6.5) | 20.4(6.0) | 20.7(5.9) | 24.7(5.8) |
| 50th Percentile (°/s) | 72.5(7.3) | 72.2(7.4) | 61.8(6.4) | 61.1(6.3) | 66.4(7.2) | 52.6(6.2) | 51.4(5.9) | 49.3(7.2) |
| 75th Percentile (°/s) | 102.3(13.1) | 101.3(13.1) | 95.8(12.0) | 94.0(12.0) | 90.1(12.1) | 88.3(12.9) | 85.8(12.3) | 70.7(8.2) |
| 90th Percentile (°/s) | 123.1(15.2) | 122.0(15.3) | 118.1(15.0) | 116.9(14.8) | 121.6(24.6) | 109.7(17.3) | 107.7(16.8) | 89.0(11.2) |
| 99th Percentile (°/s) | 153.3(17.3) | 152.8(17.0) | 148.8(17.9) | 149.0(17.7) | 191.6(53.4) | 137.1(19.1) | 135.2(18.7) | 125.1(20.4) |
| Time at low velocities (<5°/s)(%) | 0.6(0.7) | 0.6(0.7) | 2.4(2.1) | 2.2(2.1) | 0.8(0.8) | 7.9(3.8) | 7.6(3.7) | 5.7(2.0) |
| Time at high velocities (≥90°/s)(%) | 34.1(7.4) | 33.4(7.8) | 28.1(7.0) | 26.9(7.4) | 24.4(9.2) | 21.7(8.5) | 20.2(8.7) | 9.8(6.0) |
| **‘Fast’ Transfer Rate** |  |  |  |  |  |  |  |  |
| Mean (°/s) | 108.5(14.2) | 108.8(14.5) | 97.0(10.5) | 96.4(11.0) | 109.2(18.6) | 83.3(9.8) | 81.5(9.8) | 62.5(7.2) |
| 5th Percentile (°/s) | 26.2(10.3) | 28.3(10.2) | 16.8(4.5) | 18.0(5.7) | 24.5(3.7) | 7.3(1.8) | 7.5(2.6) | 5.8(1.4) |
| 10th Percentile (°/s) | 37.3(12.2) | 39.4(12.3) | 25.9(5.9) | 27.6(7.2) | 36.1(4.2) | 14.5(3.1) | 15.1(4.0) | 11.4(2.5) |
| 25th Percentile (°/s) | 66.9(14.2) | 68.2(14.3) | 52.5(8.3) | 53.1(8.9) | 60.6(5.6) | 39.1(4.9) | 39.6(5.3) | 28.7(5.3) |
| 50th Percentile (°/s) | 112.9(15.4) | 112.7(15.3) | 97.5(12.1) | 95.3(12.2) | 96.6(14.5) | 86.0(10.6) | 82.7(10.4) | 57.6(7.6) |
| 75th Percentile (°/s) | 148.4(18.8) | 147.4(19.4) | 139.3(16.4) | 137.3(17.1) | 141.9(28.2) | 125.1(17.5) | 121.5(17.3) | 89.5(10.3) |
| 90th Percentile (°/s) | 171.7(21.4) | 170.6(22.1) | 165.3(19.7) | 164.7(20.9) | 204.4(49.6) | 146.5(20.1) | 144.6(20.2) | 118.5(16.6) |
| 99th Percentile (°/s) | 207.1(23.8) | 208.0(25.0) | 201.7(23.8) | 204.0(25.0) | 310.1(77.6) | 175.5(20.9) | 174.2(21.4) | 177.8(30.5) |
| Time at low velocities (<5°/s)(%) | 0.1(0.1) | 0.1(0.1) | 0.7(0.4) | 0.7(0.4) | 0.2(0.2) | 3.6(0.9) | 3.6(0.9) | 4.6(1.3) |
| Time at high velocities (≥90°/s)(%) | 62.5(7.8) | 62.9(8.1) | 54.0(6.0) | 52.9(6.5) | 52.3(8.0) | 46.7(6.1) | 44.6(6.5) | 24.1(6.9) |

